# Genomewide Screen Identifies Peroxisomal Role in APOL1 Podocytopathy

**DOI:** 10.1101/2025.02.15.25322241

**Authors:** Jiyoung Kim, Isaac Z Karel, Huijuan Song, Megan Dewalt, Shelley Orwick, Daelynn R. Buelow, Kendyll Lee, Sergey V Brodsky, Angie Blissett, Ema Cocucci, Sharyn D. Baker, Pei-Hui Lin, Navjot S. Pabla, Sethu M Madhavan

## Abstract

The G1 and G2 variants of the APOL1 gene increase the risk of chronic kidney disease (CKD) in individuals of African descent. In the presence of secondary stressors such as inflammation and hypoxia, these gain-of-function variants can induce podocyte dysfunction and cell death through mechanisms that are not fully understood. To identify genes that influence the cytotoxic effects of APOL1 variants under hypoxic conditions, we conducted a comprehensive whole-genome RNA interference (RNAi) screen. We found that silencing several peroxisomal (PEX) genes significantly intensified the cytotoxicity associated with the G1 and G2 variants, revealing the previously unknown role of peroxisomes in APOL1-related cytotoxicity. Importantly, enhancing peroxisomal function through both genetic and pharmacological approaches led to a significant reduction in cytotoxicity linked to these variants. We also identified a peroxisomal targeting signal at the C-terminus of APOL1 that facilitates its translocation to peroxisomes during hypoxia, and mutations in this signal were found to reduce the cytotoxic effects of the variants. Collectively, our findings underscore the importance of peroxisomal function in the pathogenesis of CKD associated with APOL1 variants and suggest that targeting peroxisomes may represent an effective therapeutic strategy to mitigate CKD risk in vulnerable populations.

**TRANSLATIONAL STATEMENT:** Peroxisomal function is necessary to protect from APOL1 (kidney risk variant) KRV-mediated cytotoxicity in the presence of hypoxia, and targeting peroxisomes may represent an effective therapeutic strategy to mitigate CKD risk.

## INTRODUCTION

Variants in the APOL1 gene are associated with an increased risk of chronic kidney disease (CKD) among individuals of African ancestry.^1,2^ Under normal conditions, APOL1 expression in healthy podocytes is low; however, it can be upregulated in response to ‘second hit’ stimuli such as interferon-γ (IFN-γ) and hypoxia, resulting in podocyte dysfunction and apoptosis.^3,4^ Extensive mechanistic investigations have elucidated various pathways linking APOL1 variants to podocyte impairment, including enhanced ion channel activity, mitochondrial dysfunction, endoplasmic reticulum (ER) stress, lipotoxicity, and inflammasome activation.^5^

Not all individuals with APOL1 kidney disease risk variants (KRVs) develop CKD, indicating the potential role of modifier genes in influencing disease onset and progression. Identifying these modifier genes could facilitate the development of targeted therapeutic interventions and enhance our understanding of the mechanisms underlying APOL1 nephropathy.

In pursuit of potential disease modifier genes, we conducted an unbiased genomewide RNA interference (RNAi) screen utilizing cell culture models that mimic cell death associated with APOL1 KRVs. Our findings suggest that peroxisomal genes and the localization of APOL1 to peroxisomes are critical in mediating podocyte dysfunction related to APOL1 KRVs. This study provides the first evidence implicating peroxisomes in the pathophysiology of APOL1 nephropathy.

## METHODS

### Cell culture models of APOL1-associated cytotoxicity

HEK293 cells and human podocytes stably expressing doxycycline-inducible APOL1 variants (G0, G1, G2) were developed. Cells were cultured under hypoxia (1% O_2_) for 24-48 hours to induce cell death. Detailed methods for cell culture, RNAi induction, and pharmacological screens are available in the Supplementary Materials.

### Statistical Analysis

Data are presented as means with standard deviations and analyzed using GraphPad Prism. A p-value of <0.05 was considered significant. For two groups, a two-tailed unpaired t-test was used; for three or more groups, one-way ANOVA with Tukey’s or Dunnett’s tests was applied. No outliers were excluded, and experiments were repeated at least three times.

## RESULTS

To establish a cell culture model of APOL1 variant-induced cellular injury, we generated HEK293 cells and human podocytes with stable doxycycline-inducible expression of APOL1-G0, G1, and G2 (**Figure 1A and Supplementary Figure 1**). With comparable levels of protein expression, the cytotoxicity between APOL1-G0, G1, and G2 was not significantly different in HEK293 cells and human podocytes after 24 hours of doxycycline stimulation (**Supplementary Figure 2A-C**). Prolonged overexpression increased the cytotoxicity of APOL1-G1 and G2 compared to G0 (**Supplementary Figure 2D-F**). Importantly, in the presence of hypoxia (1% O_2_), APOL1-G1 and G2 expression led to increased cytotoxicity compared to G0 at earlier time points (24 hours) in HEK293 cells (**Figure 1B**). We used this in vitro experimental model where APOL1 overexpressing cells were challenged with a ‘second hit’ stimuli, namely hypoxia, to model APOL1 KRV-associated podocyte cell death.

**Figure 1:**
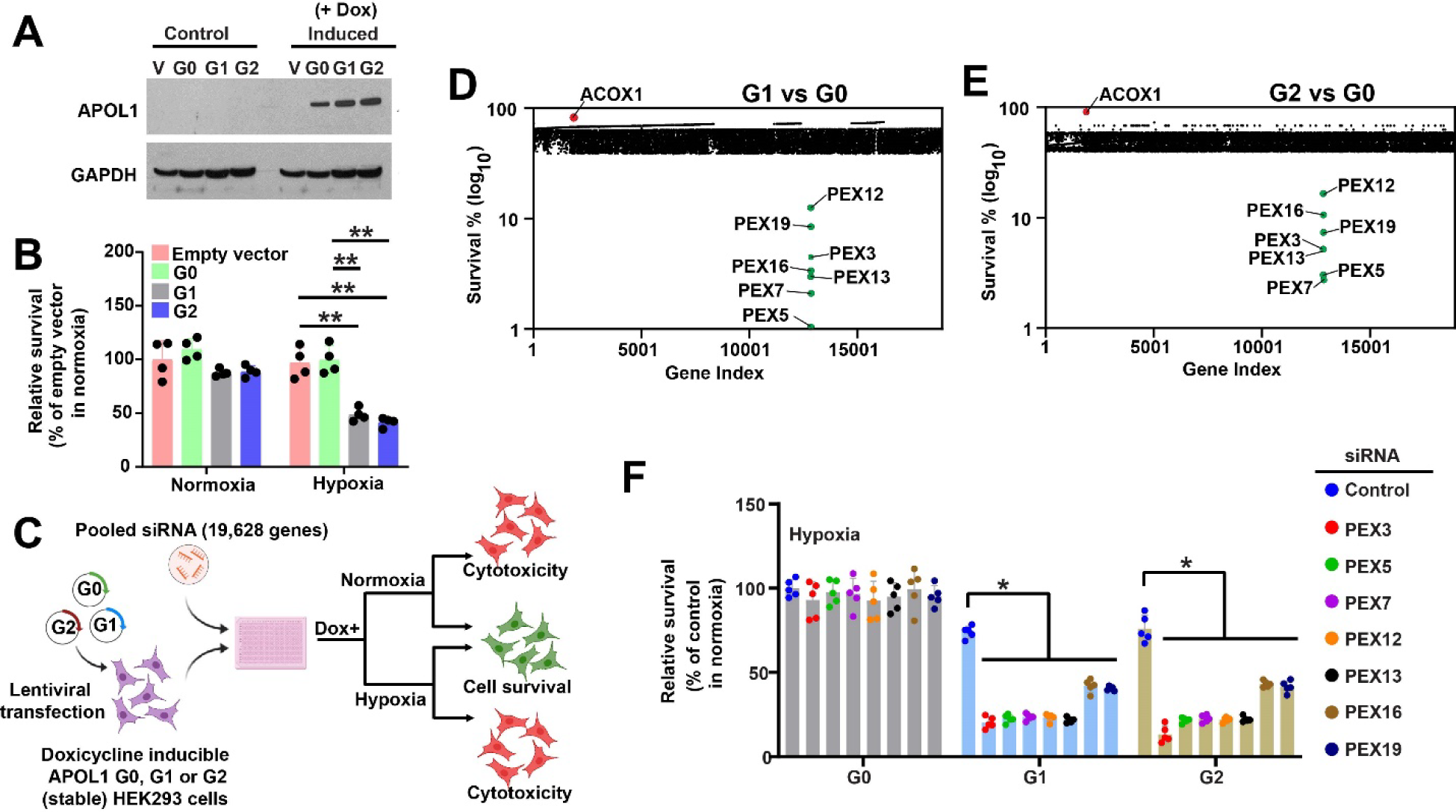
Genomewide siRNA screen identifies peroxisomes as a mediator of APOL1 variant-dependent cytotoxicity. (**A**) Western blot showing stable expression of APOL1-G0, G1, and G2 in HEK293 cells in response to doxycycline. No APOL1 expression is seen without doxycycline and with an empty vector. (**B**) APOL1 G1 and G2 expression in HEK293 cells under hypoxia (1% O2) results in increased cytotoxicity compared to G0 and empty vector control (n=4, ANOVA). (**C**) Experimental design of genome-wide siRNA screen to identify molecular pathways of APOL1 variant mediated cytotoxicity. (**D-E**) Primary siRNA screen results showing relative survival after doxycycline treatment of individual siRNA-treated genes comparing G0 to G1 (D) and G0 to G2 (E) in hypoxia. Experiments performed in triplicates. (**F**) Validation of primary hits by separate siRNAs in HEK293 cells. In the presence of hypoxia, knockdown of individual peroxisomal genes in HEK293 cells expressing empty vector or APOL1-G0 does not increase cytotoxicity. siRNA knockdown of PEX genes increased G1 and G2-associated cytotoxicity. Bar graphs represent five biologically independent replicates with results expressed as mean ± SD. (**p<0.001, *p<0.05).

We conducted a genome-scale siRNA screen targeting 19,628 genes to identify genetic factors influencing cytotoxicity induced by APOL1-KRVs under hypoxic conditions (**Figure 1C and Supplementary Figure 3A**). This screening strategy was informed by the concept of ‘synthetic lethality’ as applied in cancer biology, a method designed to identify genes whose knockdown synergistically compromises cell survival. Our goal was to pinpoint specific genes that either potentiate or mitigate the cytotoxic effects associated with the APOL1-G1 and G2 variants. The experimental procedures for the primary siRNA screen, as well as the subsequent data analysis, were rigorously conducted using previously described methods from our group.^6,7^

Surprisingly, our results revealed that the knockdown of genes associated with peroxisomal biogenesis—specifically PEX genes (*PEX3, PEX5, PEX7, PEX12, PEX13, PEX16, PEX19*)—led to an increase in APOL1-G1 and G2-mediated cytotoxicity. In contrast, silencing acyl-CoA oxidase 1 (*ACOX1*), a peroxisome-localized enzyme, resulted in a decrease in cytotoxicity compared to the control condition (G0) (**Figure 1D-E and Supplementary File 1**). To ensure the reliability of these findings, we performed rigorous secondary validation using different siRNAs from Sigma Aldrich and cell viability assays (**Figure 1F**). The secondary screen confirmed the initial findings from the primary screen (**Figure 1F** and **Supplementary Figure 3**), indicating a critical role for peroxisomes in the mechanism of APOL1 KRV-mediated cell death.

Based on our screening results, we hypothesized that enhancing peroxisomal function could reduce cell death mediated by APOL1 KRV. To investigate this, we employed both genetic and pharmacological approaches to modulate peroxisome function in our cell culture model. Overexpressing PEX3 effectively mitigated the cytotoxic effects of APOL1-G1 and G2 under hypoxic conditions (**Figure 2A-B**). Several PPAR-α agonists, which are known to boost peroxisomal function, also alleviated the cytotoxicity caused by APOL1-G1 and G2 in hypoxia (**Figure 2C**). Notably, pemafibrate, a selective PPAR-α modulator with significantly higher potency—approximately 2500 times greater than traditional agents like fenofibrate— demonstrated the most effective profile in reducing cytotoxicity associated with the APOL1-G1 and G2 variants (**Figure 2D-E**). To assess whether APOL1 KRVs impact peroxisomal function, we measured catalase activity in isolated peroxisomes. Interestingly, peroxisomes from HEK293 cells expressing APOL1-G1 and G2 exhibited higher catalase activity compared to those expressing G0 (**Supplementary Figure 4**). The elevated catalase activity induced by APOL1 G1 and G2 was reversed by overexpression of PEX3 and treatment with pemafibrate (**Figure 2F-G**).

**Figure 2:**
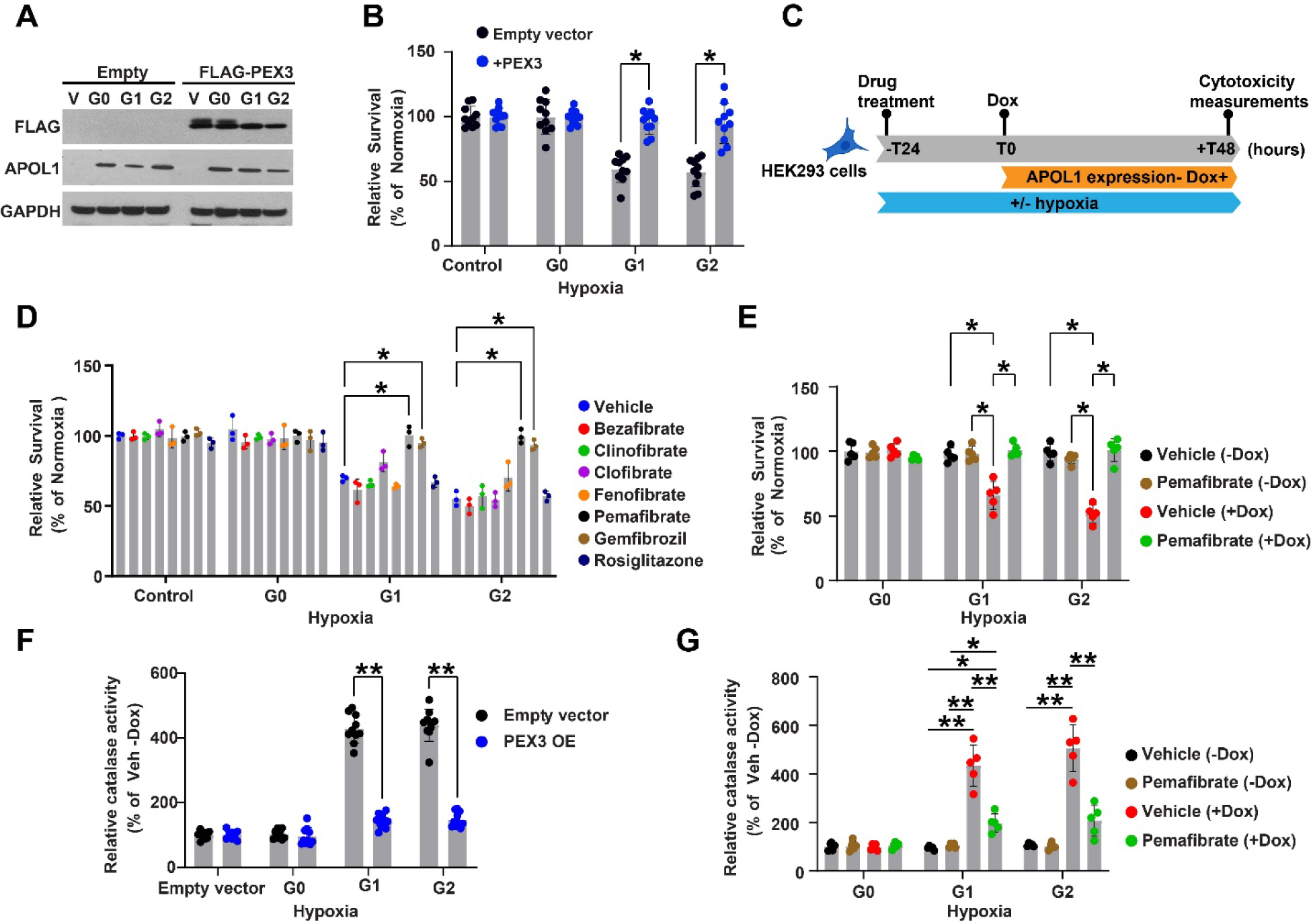
Genetic or pharmacological improvement of peroxisome function mitigates APOL1-associated cellular dysfunction. (**A**) Western blot showing overexpression of PEX3 in HEK293 cells stably expressing doxycycline-inducible APOL1-G0, G1 or G2. (**B**) Overexpression of PEX3 in HEK293 cells stably expressing APOL1s rescues the cytotoxicity caused by G1 and G2 under hypoxic conditions. (n=4), (*p<0.001). (**C**) HEK293 cells stably transfected with APOL1 G0, G1, or G2, were pre-treated with drugs for 24 hours, followed by induction of APOL1s expression with doxycycline. Cell viability was measured 48 hours after doxycycline induction. (**D**) Screening of PPAR modulating drugs showing effect on mitigating the cytotoxicity of APOL1 G1 and G2 variants in HEK293 cells. Cells were treated with drugs for the entire 72 hours of the experiment. These results show that genetic or pharmacological augmentation of peroxisomal function can mitigate G1 and G2-associated cytotoxicity (two experimental replicates, each with n=3). Cell survival was measured relative to normoxia. (*p<0.001). (**E**) Pemafibrate improves the survival of APOL1-G1 and G2 under hypoxia (n=5). (*p<0.001). (**F-G**) PEX3 overexpression and pemafibrate treatment reduced catalase activity in peroxisomes isolated from HEK293 cells expressing APOL1 G1 and G2. (**p<0.001, *p<0.05).

APOL1 is known to localize to various intracellular organelles, including the endoplasmic reticulum (ER), but its presence in peroxisomes has not been investigated. So, we isolated peroxisomes from HEK293 cells and human podocytes and found no APOL1 localization in peroxisomes under normal conditions. However, hypoxia triggered APOL1 localization to peroxisomes (**Figure 3A**). Protein transport to peroxisomes is driven by peroxisomal targeting signals (PTS). The canonical PTS-1 is characterized by a C-terminal SKL sequence. In our study, we identified a -SKHL- sequence at the C-terminus of APOL1 and explored its potential as a PTS (**Figure 3B**). We created mutants by changing SKHL to SVHL and transfected them into HEK293 cells. The peroxisomes in cells expressing APOL1-(G0, G1, G2)-SVHL exhibited reduced localization to peroxisomes under hypoxic conditions compared to the reference variants (**Figure 3C**). This mutation did not influence ER localization (**Figure 3C**). The reduced peroxisomal localization was associated with decreased cytotoxicity (**Figure 3D**) and lower peroxisomal catalase activity linked to APOL1-G1 and G2 (**Figure 3E**). Notably, while G0, G1, and G2 all localized to peroxisomes under hypoxia, only G1 and G2 enhanced catalase activity. These findings provide direct evidence of the role of peroxisomal targeting in the cytotoxicity of APOL1 KRVs.

**Figure 3:**
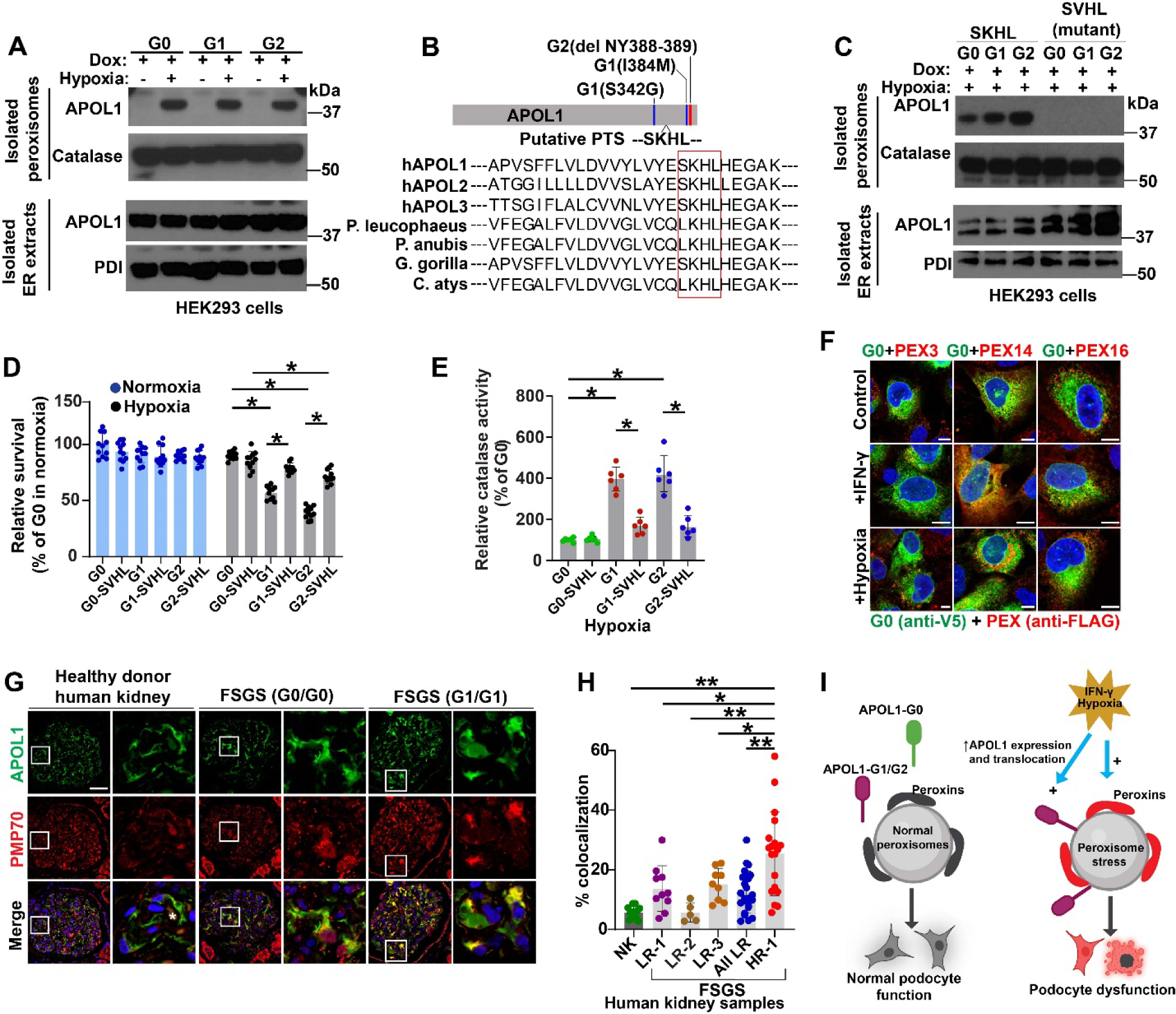
APOL1 interacts with peroxisomes and causes peroxisomal stress in response to hypoxia. (**A**) Western blot showing APOL1 G0, G1, and G2 translocation to peroxisomes isolated from HEK293 cells and human podocytes in response to hypoxia. Localization to ER remained unchanged. (**B**) Schematic representation of putative peroxisome localization signal (PTS) in APOL1 C-terminus and location of G1 and G2 variants. The sequence alignment in the C-terminal region carrying the putative PTS region is shown. (**C**) Western blot for APOL1 in isolated peroxisomes from HEK293 cells shows a reduction in the localization of APOL1-SVHL mutants to peroxisomes in response to hypoxia compared to the non-mutant version. APOL1-SKHL mutations did not influence localization to ER. (**D**) PTS influences the cytotoxicity of APOL1 G1 and G2 variants. Mutation of putative peroxisomal targeting signal (PTS) in APOL1 C-terminus reduced the cytotoxicity of APOL1 G1 and G2 in response to hypoxia. (n=11 in 3 biological replicates). (*p<0.001). (**E**) Loss of PTS results in reduced catalase activity in HEK293 cells expressing G1 and G2 in hypoxia. (n=6), (*p<0.001). (**F**) Confocal immunofluorescence microscopy showing expression and colocalization of APOL1-G0 with PEX3, PEX14, and PEX16 in differentiated human podocytes in the presence and absence of IFN-γ and hypoxia. Scale bars: 10 µm (**G**) Confocal immunofluorescence microscopy showing colocalization of APOL1 with peroxisomal membrane protein PMP70 in healthy human kidneys and FSGS biopsies with and without high-risk *APOL1* genotype. Capillary loops denoted by asterisk. Scale bars: 20 µm (**H**) Pixel-based analysis of APOL1 and PMP70 expression in glomeruli shows increased colocalization in FSGS patients carrying APOL1-HR than LR genotype and healthy human kidneys. (**p<0.001, *p<0.05). (**I**) Model for peroxisome dysfunction mediating APOL1 KRVs associated CKD pathogenesis. APOL1-G0, -G1, or -G2 expression in podocytes in the absence of peroxisomal dysfunction does not result in cytotoxicity. Increased expression of APOL1-KRVs and subsequent translocation to peroxisomes in the presence of second stress can mediate podocyte dysfunction by dysregulating peroxisomal function.

In human podocytes, APOL1-G1 and G2 expression also caused increased cytotoxicity and catalase activity in the presence of hypoxia (**Supplementary Figure 5A-B**). Confocal immunofluorescence microscopy showed colocalization of APOL1-G0 with PEX3, PEX14, and PEX16 with IFN-γ and hypoxia in human podocytes (**Figure 3F and Supplementary Figure 6A-B**). We then sought to corroborate these results with STORM imaging. In human podocytes stably co-expressing APOL1-G0 and PEX14, APOL1-G0 partially colocalized with PEX14 at baseline (**Supplementary Figure 6C**) and increased in response to IFN-γ and hypoxia (**Supplementary Figure 6D**). Next, we used a proximity ligation-based pulldown strategy to detect if APOL1 interacted with the peroxisomal membrane. Human podocytes stably expressing APOL1-G0 fused to turboID, a biotin ligase enzyme that biotinylated proteins in proximity in the presence of biotin, were generated (**Supplementary Figure 7A**). We identified that APOL1-G0 interacted with the peroxisomal membrane as detected by proximity ligation of PEX14 in addition to ER and mitochondrial matrix proteins (**Supplementary Figure 7B-C**). Finally, we sought to test the APOL1 localization to the peroxisomes in human kidneys. Confocal immunofluorescence microscopy showed colocalization of APOL1 with peroxisomal membrane proteins, PMP70, and PEX14 in podocytes of healthy human kidneys (**Figure 3G and Supplementary Figure 8A**). Biopsies from patients with focal segmental glomerulosclerosis (FSGS) carrying the APOL1-HR genotype showed increased colocalization of APOL1 with PMP70 in glomeruli (**Figure 3G-H**). Glomerular expression of peroxisomal proteins identified by mass spectrometry (LC-MS) differentiated FSGS from healthy kidneys (**Supplementary Figure 9A**). Finally, the glomeruli of patients with FSGS showed reduced expression of peroxisomal biogenesis proteins PEX5, PEX14, and PEX19 using LC-MS studies (**Supplementary Figure 9B**).

## DISCUSSION

The G1 and G2 risk alleles of APOL1 are critical contributors to FSGS, hypertension-associated renal failure, and HIV nephropathy, functioning as toxic gain-of-function variants.^8^ While the correlation with renal dysfunction is well-established, the specific cellular contexts and mechanistic pathways remain insufficiently characterized. Initially recognized for its protective role against *Trypanosoma* species, APOL1’s trypanolytic activity is mediated through its ion channel function, with the activation of detrimental channel properties proposed to initiate renal injury.^9,10^ Current evidence suggests that aberrant channel function in podocytes, where APOL1 is predominantly localized to the endoplasmic reticulum and plasma membrane, underpins this pathology.^11,12^ This study identifies a peroxisomal localization signal within APOL1, proposing that peroxisomal dysfunction represents a previously unrecognized element in the pathogenesis of APOL1 nephropathy (**Figure 3I**).

Peroxisomes are single-membrane-bound organelles essential for fatty acid oxidation and cellular redox balance, containing over 50 species- and cell-type-specific enzymes.^13^ They are involved in metabolic processes such as β-oxidation of very long-chain fatty acids (VLCFAs) and hydrogen peroxide detoxification. Dysregulation of peroxisomal function, particularly in fatty acid oxidation, is implicated in acute kidney injury (AKI) and chronic kidney disease (CKD), yet their role in APOL1 nephropathy remains unexplored. Present in all superphyla of eukaryotes; peroxisomes exhibit homologous features in biogenesis, function, and morphology despite their diversity. In trypanosomes, specialized peroxisomes known as glycosomes can contain over 90% of their protein content as glycolytic enzymes, particularly in *Trypanosoma brucei* residing in the mammalian bloodstream.^14^ We hypothesize that the trypanolytic activity of APOL1 may depend on peroxisomal localization, warranting future experimental investigation.

The import of peroxisomal proteins is initiated by the recognition of a peroxisomal-targeting signal (PTS), typically characterized by a sequence motif at the C-terminus (PTS1) or one near the N-terminus (PTS2).^15,16^ In this study, we identified an atypical PTS1 site near the C-terminus of APOL1. Notably, under standard conditions, the peroxisomal localization of G0, G1, and G2 variants was minimal; however, hypoxia-induced peroxisomal localization of all APOL1 variants. Mutation of a positively charged lysine residue within the PTS1 sequence abolished this localization. Importantly, hypoxia-induced cytotoxicity associated with G1 and G2 variants was also negated upon mutation of the PTS1 site. The SKHL to SVHL mutation did not affect protein stability or localization to the endoplasmic reticulum. Additionally, the hypoxia-induced translocation of APOL1 to peroxisomes correlated with increased catalase activity, a phenotype that was suppressed by the SKHL to SVHL mutation. Collectively, these findings provide direct evidence that APOL1-associated cytotoxicity is at least partially dependent on its peroxisomal localization.

Not all individuals harboring APOL1 kidney risk variants (KRVs) manifest kidney dysfunction, indicating the potential involvement of an unidentified “second hit” as well as other genetic or environmental modifiers. Through a genomewide screening methodology, we identified critical genes necessary for the survival of cells expressing APOL1-G1 and G2 variants under hypoxic conditions, a recently recognized potential second hit. Importantly, the knockdown of peroxisomal genes did not affect the viability of cells with the normal APOL1-G0 genotype; however, cells with G1 and G2 variants displayed increased susceptibility to hypoxia-induced cell death. This suggests that impaired peroxisomal function may exacerbate APOL1 nephropathy. This phenomenon mirrors synthetic lethality observed in cancer biology, where mutant cells rely on specific genes for survival, thereby facilitating targeted therapeutic approaches.^17^ Consequently, in addition to employing small molecule inhibitors of APOL1^18^, the augmentation of peroxisomal function through PPARα agonists could potentially attenuate disease progression. Future studies are also warranted to determine whether individuals with APOL1 KRVs receiving PPARα agonist treatment exhibit reduced susceptibility to kidney disease.

Based on previous studies, the gain-of-function of APOL1 KRVs causes increased ion channel activity, mitochondrial dysfunction, ER stress, and lipotoxicity, leading to podocyte dysfunction.^5^ By the structural and functional cross-talk of peroxisomes with these organelles^19-23^, dysregulated peroxisomal function caused by APOL1 KRVs can lead to these observed cellular phenotypes. The link between APOL1 and peroxisomal function that we have identified is based on *in vitro* studies. Future studies using transgenic mice with peroxisomal targeting mutants of APOL1 will confirm the direct effect of APOL1 KRVs on peroxisomal dysfunction leading to kidney disease. Additionally, the presence of variants in peroxisomal biogenesis and functional genes that could modify kidney disease predisposition in individuals with APOL1 KRVs needs to be studied.

## Data Availability

All data produced in the present study are available upon reasonable request to the authors

## ACKNOWLEDGEMENTS

This work was supported by funds from The Ohio State University (OSU) Department of Internal Medicine and National Institutes of Health grants R01DK132230 (NSP), and K08DK123411 (SMM).

## SUPPLEMENTARY DATA

## METHODS

### Plasmid constructs and cell lines

For the generation of stable cell lines, the following plasmid constructs were generated. V5 tags were introduced in the APOL1 G0, G1, or G2 sequence between the signal peptide (SP: aa 1-30) and the rest of the cDNA. We used this strategy because introducing tags to the N-terminus before the signal peptide resulted in the cleavage of the tag along with the signal peptide. For generating all the plasmids, we utilized APOL1-G0, -G1, and -G2 in the naturally occurring haplotype background (G0 carrying K150, I228, K255, and G1 and G2 with E150, I228, and K255). For lentiviral plasmid generation, all constructs were cloned into pCW57.1 (Addgene #41393) using the AgeI and NheI sites. For transient transfection of HEK293 cells SP-FLAG-APOL1 cDNA was cloned into pCMCTNT vector (Promega) using the EcoRI and NotI sites. Immortalized human podocytes were cultured and differentiated as previously described.^24^ HEK293 and podocyte stable cell lines were generated using lentiviral transfection and selected using puromycin. Human podocytes were grown in RPMI 1640 media with 1X insulin-transferrin-selenium (ITS-G) (Thermo Scientific), and HEK293 cells were grown in DMEM media with both media containing 10 % tetracycline-free FBS (Takara). The plasmids and cell lines generated in the study are summarized in **Supplementary Table 1**.

### Genomewide RNAi screening

RNAi screen and secondary validation were performed using methods described in our previous studies.^6,7^ HEK293 cells stably expressing APOL1-G0, G1, and G2 under doxycycline regulation were plated in 384-well plates to achieve 50% confluence in Dulbecco’s Modified Eagle Medium (DMEM) supplemented with 10% fetal bovine serum under normoxic conditions. During plating, the cells underwent reverse transfection with either control small interfering RNA (siRNA) or a comprehensive siRNA library (Dharmacon, Human ON-TARGETplus siRNA Library - Whole Genome), which targets 19,628 genes with four distinct pooled siRNAs per gene. Lipofectamine RNAiMAX reagent (Life Technologies) was utilized to transiently transfect the siRNAs. Following a 24-hour incubation period post-transfection, APOL1 gene expression was induced by replacing the medium with doxycycline-containing DMEM with 10% fetal bovine serum. Subsequently, cells were transferred to a hypoxia chamber, maintained at 1% oxygen, for an additional 24 hours. Cellular viability was assessed using the CellTiter-Glo assay (Promega). The primary screening was conducted in triplicate, and data analysis was performed following previously established methodologies.^6,7^ For secondary validation, similar experiments were conducted in 96 well plates utilizing distinct siRNAs (Sigma, Mission Predesigned) targeting peroxisome-related genes.

### Pharmacological screening

HEK293 cells stably expressing APOL1-G0, G1, and G2 under doxycycline regulation were seeded in 96-well plates to achieve 50% confluence in Dulbecco’s Modified Eagle Medium (DMEM) supplemented with 10% fetal bovine serum under normoxic conditions. Following a 24-hour incubation, the cells were treated with either dimethyl sulfoxide (DMSO) or a 1 µM concentration of PPARα agonists for 24 hours, with the agonists retained for subsequent experimental procedures. APOL1 gene expression was then induced by replacing the culture medium with doxycycline-containing DMEM supplemented with 10% fetal bovine serum. The cells were subsequently transferred to a hypoxia chamber maintained at 1% oxygen for an additional 24 hours. Cellular viability was assessed using both the CellTiter-Glo assay (Promega) and the trypan blue exclusion method.

### Gene expression analysis

Total RNA (1 µg) from HEK293 cells was reverse transcribed using the RevertAid First Strand cDNA Synthesis Kit (Thermo Fisher Scientific). Quantitative reverse transcription polymerase chain reaction (qRT-PCR) was subsequently conducted on a QuantStudio 7 Flex Real-Time PCR System (Thermo Fisher Scientific), employing SYBR Green Master Mix and pre-designed gene-specific primers for PEX genes (Sigma). Relative gene expression was calculated using the comparative CT (ΔΔCT) method, with β-actin serving as the reference gene.

### Cytotoxicity assays

Cytotoxicity assays in response to APOL1 expression in HEK293 cells and human podocytes were performed using trypan blue exclusion, CytoTox-ONE LDH assay (Promega), Multiflour MultiTox-Fluor Multiplex cytotoxicity assay (Promega) and clonogenic survival assays.

<u>Clonogenic survival assay:</u> Human podocytes stably expressing APOL1-G0, G1, and G2 under the control of doxycycline were plated 6-well plates at 400 cells/well density. APOL1 expression was induced with doxycycline (200 ng/ml), and media was changed every 48 hours and cultured for 12 days. Cells were fixed with 6% glutaraldehyde and stained with 1% crystal violet. Colonies were counted, and surviving fractions were calculated.^25^ <u>LDH release assay:</u> Human podocytes with stable doxycycline-inducible expression of APOL1-G0, G1, and G2 were plated in 96-well plates (40,000 cells/well). Twenty-four hours after plating, APOL1 expression was induced with doxycycline (200 ng/ml). LDH release as a measure of cytotoxicity was measured using CytoTox-ONE Homogenous Membrane Integrity Assay (Promega) at 48 and 72 hours as per manufacturer’s instructions. The assays were performed in three independent experiments (n=3), with each experiment in triplicates (n=9). Cytotoxicity (%) was calculated as (experimental LDH release-media background/maximal LDH release-media background). <u>MultiTox-Fluor Multiplex cytotoxicity assay (Promega):</u> Cells were plated to 96-well plates (100,000 cell/well for cytotoxicity measurements at 8, 16, 24 hours, and 40,000 cells/well for cytotoxicity measurements at 48 and 72 hrs). The cytotoxicity and cell viability measurements were performed per the manufacturer’s instructions at 8, 16, 24, 48, and 72 hours after doxycycline stimulation (200 ng/ml). Groups were compared by ANOVA followed by post-hoc multiple comparison tests, and p-value less than 0.05 was considered significant. <u>Trypan blue exclusion assay:</u> Cell viability was further evaluated utilizing the trypan blue exclusion assay using previously described methods.^6^ In brief, cells were harvested and stained with trypan blue, followed by quantification using a hemocytometer and/or the Countess Automated Cell Counter (Thermo Fisher). Translucent cells were identified as viable, whereas blue-stained cells were considered non-viable. The viability percentage was determined by calculating the ratio of viable cells to the total cell count, with each sample processed in triplicate.

### Organelle fractionation and catalase assay

Peroxisomes and endoplasmic reticulum (ER) fractions were isolated from cultured cells utilizing commercially available kits (Sigma, PEROX1-1KT, and ER0100-1KT), in accordance with the manufacturer’s protocols. The purity of the isolated peroxisomes and ER fractions was confirmed via western blot analysis employing organelle-specific markers. Catalase activity was assessed using a kit from Cayman Chemicals (707002), with the results normalized to protein concentrations determined by the BCA assay (Pierce).

### Proximity ligation and pulldown

Human podocytes stably expressing V5-tagged APOL1-G0 fused to turboID (**Supplementary Table 1**) were grown in differentiating conditions. APOL1-G0-turboID expression was induced with doxycycline (200 ng/ml) for 16 hours. Biotinylation of proteins proximal to APOL1-G0 was performed by the addition of 200 µM biotin for one hour. Cells were lysed with lysis buffer containing 50 mM Tris-HCl pH 8, 150 mM sodium chloride,1% IPEGAL CA-630, 0.25% sodium deoxycholate, and centrifuged at 10,000 g for 5 minutes at 4^0^C. The supernatant was incubated with streptavidin magnetic beads (Thermo) for one hour at 4^0^C. Beads were washed with lysis buffer, and proteins bound to the beads were eluted by heating in 4X Laemmli buffer (Bio-Rad Laboratories, Hercules, CA) supplemented with 20 mM DTT and 2 mM biotin, followed by Western blot to detect bound biotinylated proteins.

### Western blots

For western blots, cells were lysed using lysis buffer containing 50 mM Tris, pH 7.5, 150 mM NaCl, 1% NP40, 0.5% sodium deoxycholate. Samples were heated in Lamelli buffer and were resolved using Bis-Tris gel and transferred to PVDF membranes. The membrane was blocked using 5% non-fat dry milk and incubated overnight with primary antibodies (**Supplementary Table 2**) and appropriate secondary antibodies. Chemiluminescence was detected using SuperSignal West Pico PLUS chemiluminescent substrate (ThermoScientific).

### Confocal immunofluorescence and STORM microscopy

Cells grown on coverslips were fixed with 4% paraformaldehyde (PFA) at room temperature for 15 minutes, washed in PBS, and permeabilized with 3% BSA with 0.2% Triton-X in PBS at room temperature for 20 minutes. Cells were then incubated with primary antibodies overnight at 4^0^C (**Supplementary Table 2**) and secondary antibodies for 2 hours at room temperature in blocking buffer. Formalin-fixed paraffin-embedded tissue sections were rehydrated in ethanol gradient followed by heat-induced antigen retrieval using citrate (10 mM sodium citrate, 0.05% Tween-20, pH 6) or Tris-EDTA (10 mM Tris-base, 1 mM EDTA, 0.05% Tween-20, pH 9) buffer. Sections were blocked with 10% serum in PBS-0.2% Tween-20 for 1 hour at room temperature and incubated with primary antibodies overnight at 4°C followed by appropriate secondary antibodies for one hour at room temperature. Confocal images were obtained using an Olympus FV3000 confocal microscope. STORM images were captured from cultured human podocytes using Nikon N-STORM microscope.

### Human samples

Healthy human kidney samples were obtained from biopsies of transplant donor kidneys (n=4), and biopsies from patients with FSGS (n=12) were obtained from OSU biorepository. The human subject studies that were performed were approved by the institutional review board at Ohio State University.

### Mass spectrometry and proteomics studies

Healthy donor kidneys and FSGS biopsies in formalin-fixed paraffin-embedded blocks were cut (10 μm) and mounted onto PEN membrane slides (Zeiss). After removing the paraffin and rehydrating the slides through a series of ethanol gradients, the glomeruli were isolated using laser capture microdissection (PALM Technologies, Carl Zeiss Microimaging GmbH, Munich, Germany). The dissected tissue sections were then captured into a solution containing 0.5% Rapigest (Waters Corporation, Milford, MA) in 50 mM sodium bicarbonate. Subsequently, for protein extraction, the samples were boiled 20 minutes, followed by a 2-hour incubation in a 60°C water bath. Protein digestion was facilitated by the addition of trypsin (Promega, Madison, WI) at a ratio of 1:30 (trypsin), based on an estimated recovery of 2 μg of protein from 10,000 cells. The protein-trypsin mixture was left to incubate overnight at 37°C, followed by treatment with formic acid at a final concentration of 30-40% for 30 minutes at room temperature to precipitate the Rapigest completely. After centrifugation at 15,000 rpm at 4°C for 10 minutes to remove cell debris and precipitated Rapigest, the samples were dried using a speedvac. The dried peptides were then solubilized by resuspending in 2% acetonitrile with 0.1% formic acid.

Liquid chromatography tandem-mass spectrometry (LC-MS/MS) analysis was conducted using a Thermo Scientific Easy1200 nLC coupled with a tribrid Orbitrap Eclipse mass spectrometer (Thermo Scientific, Waltham, MA). In the process, inline de-salting was achieved through a reversed-phase trap column (100 μm × 20 mm) packed with Magic C18AQ (5-μm 200Å resin) followed by peptide separations on a reversed-phase column (75 μm × 270 mm) packed with ReproSil-Pur C18AQ (3-μm 120Å resin), directly mounted on the electrospray ion source. A 180-minute gradient was employed, utilizing a two-mobile-phase system consisting of 0.1% formic acid in water (A) and 80% acetonitrile in 0.1% formic acid in water (B). This gradient spanned from 8% to 30% B over 180 minutes, followed by specific increments over defined periods, reaching 90% B for a subsequent duration. The chromatographic separation was conducted at a flow rate of 300 nL/minute. A spray voltage of 2300 V was applied to the electrospray tip, while a FAIMS source with varied compensation voltages (-40, -60, -80) was utilized. The Orbitrap Eclipse instrument operated in data-dependent mode, with MS survey scans conducted in the Orbitrap, while MS/MS spectra acquisition was performed in the linear ion trap using HCD activation. Selected ions were dynamically excluded for 60 seconds after a repeat count of 1.

Raw MS data underwent processing using Proteome Discoverer (v2.5.0.400, Thermo Fisher Scientific). Spectra were searched against the human proteome obtained from Uniprot (August 2022) using Sequest HT, applying fixed modification of carbamidomethylation of cysteine and variable modifications of oxidation of methionine, oxidation of proline, and N-terminal acetylation. Tryptic peptides were allowed with specified maximums, and precursor and fragment ion tolerances were set at 10 ppm and 0.6 Da, respectively. False discovery rates (FDRs) for both peptide and protein identification were rigorously set at 0.01.

### Supplementary Files

**Supplementary File 1:** Cytotoxicity data from genomewide siRNA screens done in HEK293 cells expressing APOL1 G0 vs G1 or G2.

## Supplementary Figures

**Supplementary Figure 1:**
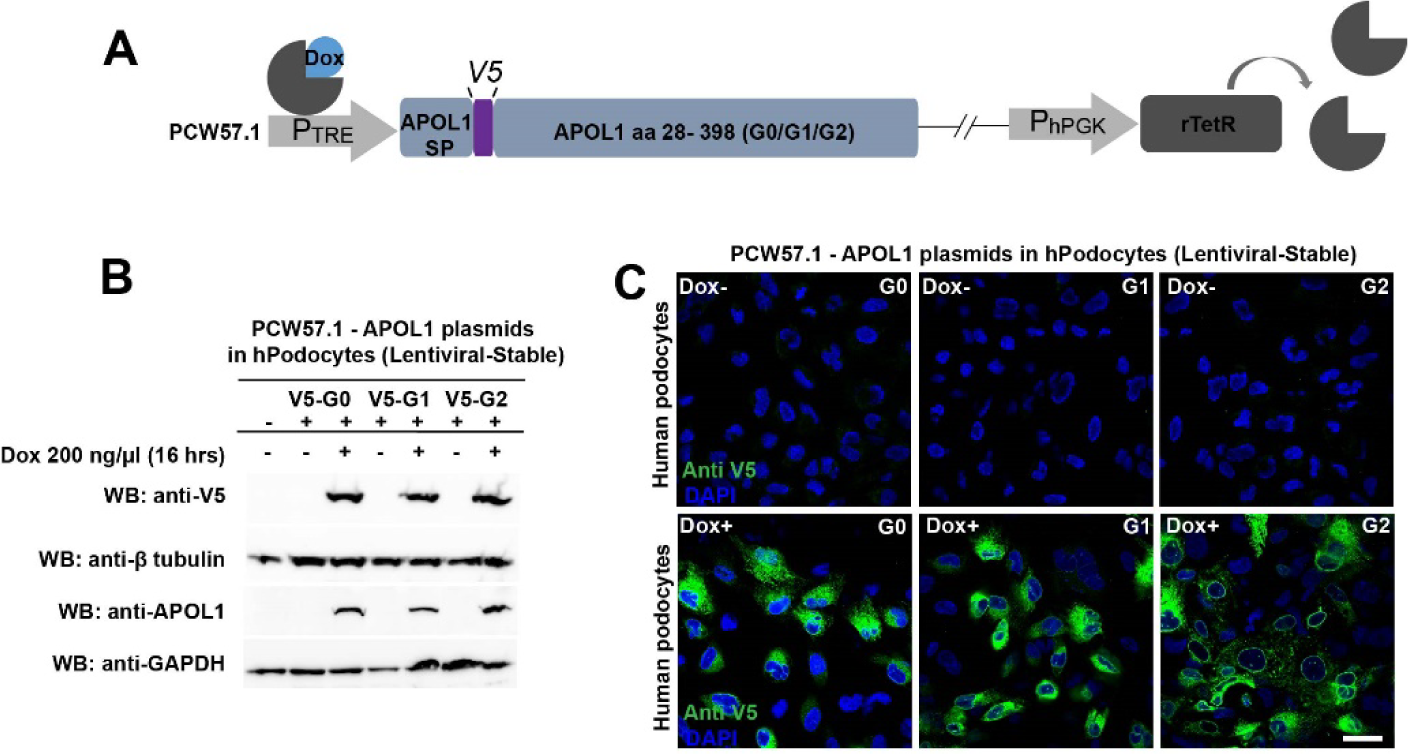
Generation and characterization of HEK293 and human podocytes with stable inducible APOL1-G0, G1, and G2 expression. **(A)** Schematic representation of APOL1-G0, G1, or G2 vector used to generate stable cell lines. V5 tag is inserted into the sequence after the signal peptide to avoid cleavage along with the signal peptide. APOL1 expression by pTRE promoter is under the control of doxycycline. **(B)** Western blot for V5 and APOL1 in human podocytes following stable lentiviral transfection expressing V5-APOL1-G0, G1, or G2 induced by doxycycline. **(C)** Confocal immunofluorescence microscopy showing expression of V5-APOL1-G0, G1, or G2 in the absence and presence of doxycycline in human podocytes. Scale bars: 50 µm.

**Supplementary Figure 2:**
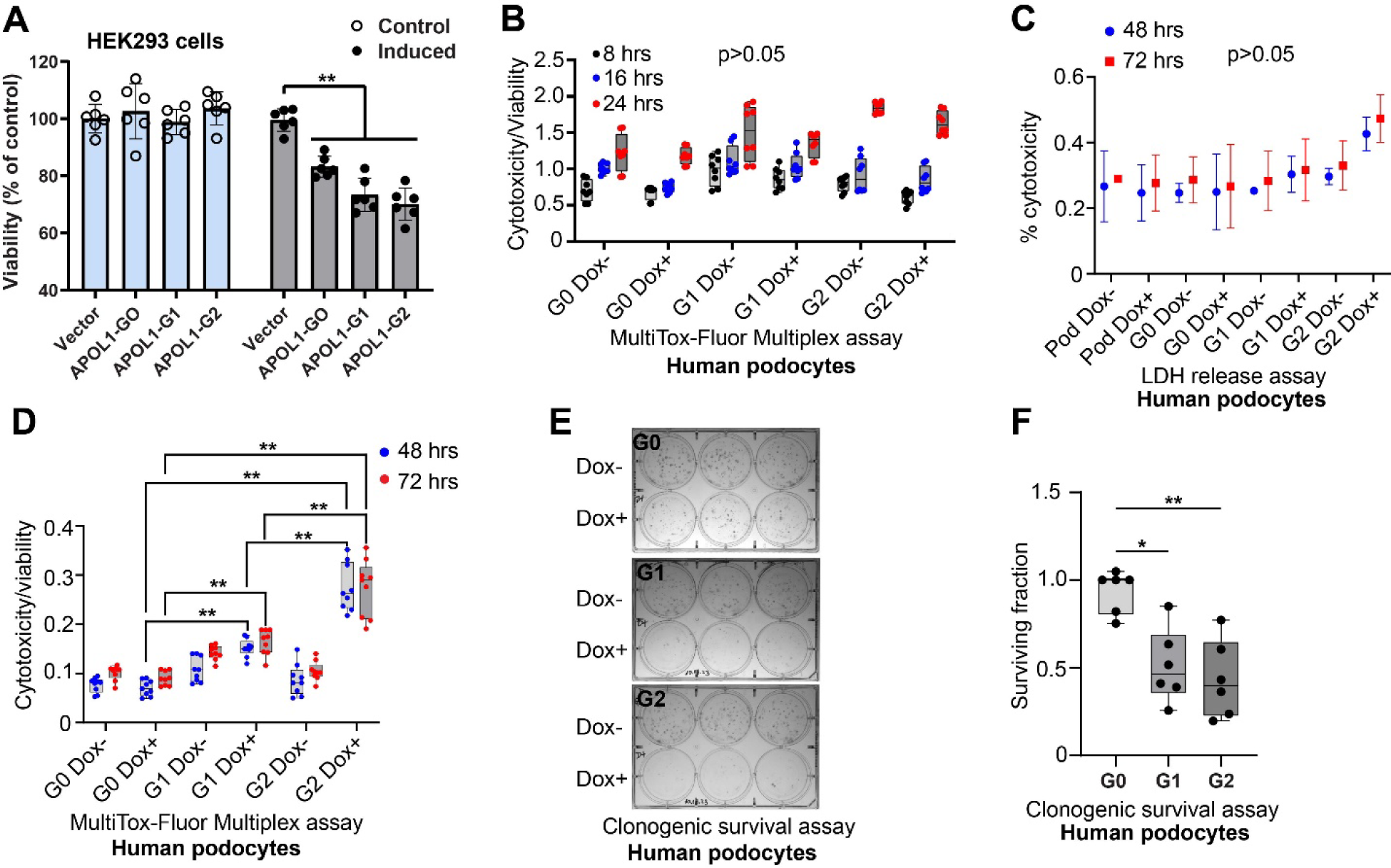
Characterization of cytotoxicity in HEK293 and human podocytes with stable inducible APOL1-G0, G1, and G2 expression. (**A**) APOL1 G0, G1 or G2 stable expression in HEK293 cells by doxycycline induction does not result in differential cytotoxicity at 24 hours (trypan blue exclusion assay) (n=6). (**C**) Human podocytes with stable doxycycline-inducible expression of APOL1-G0, G1, and G2 were plated in 96-well plates (100,000 cells/well). APOL1s expression was induced with doxycycline after 24 hours and cytotoxicity was measured at 8, 16 and 24 hours using MultiTox-Fluor Multiplex cytotoxicity assay (Promega). Expression of APOL1 G0, G1, and G2 does not result in differential cytotoxicity at 8, 16, or 24 hours (n= 8). p not significant. (**F**) LDH release as a measure of cytotoxicity was measured using CytoTox-ONE Homogenous Membrane Integrity Assay (Promega) at 48 and 72 hours after doxycycline induction of APOL1s in human podocytes. The assays were performed in three independent experiments (n=3), with each experiment in triplicates (n=9). p not significant. (**D**) MultiTox-Fluor Multiplex cytotoxicity assay (Promega) measuring cytotoxicity and cell viability in human podocytes at 48 and 72 hours after doxycycline induction of APOL1s (n= 8). (**E-F**) Clonogenic survival assay in human podocytes stably expressing APOL1-G0, G1, and G2 under the control of doxycycline. Two separate experiments with three biological replicates were done. (**p<0.001, *p<0.05).

**Supplementary Figure 3:**
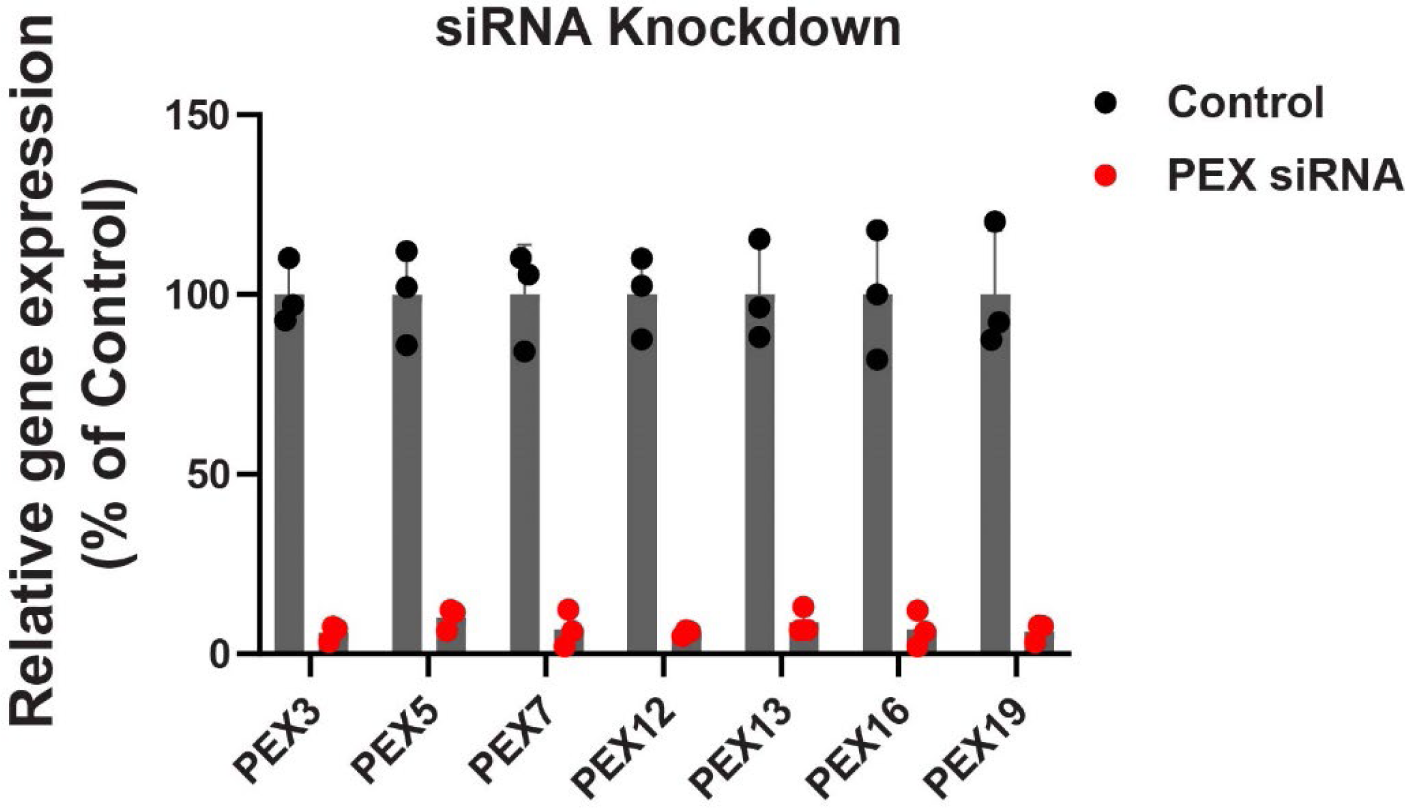
siRNA knockdown of *PEX* genes. (**A**) qPCR showing knock down of PEX genes in HEK293 cells used for secondary validation of whole-genome siRNA screen.

**Supplementary Figure 4:**
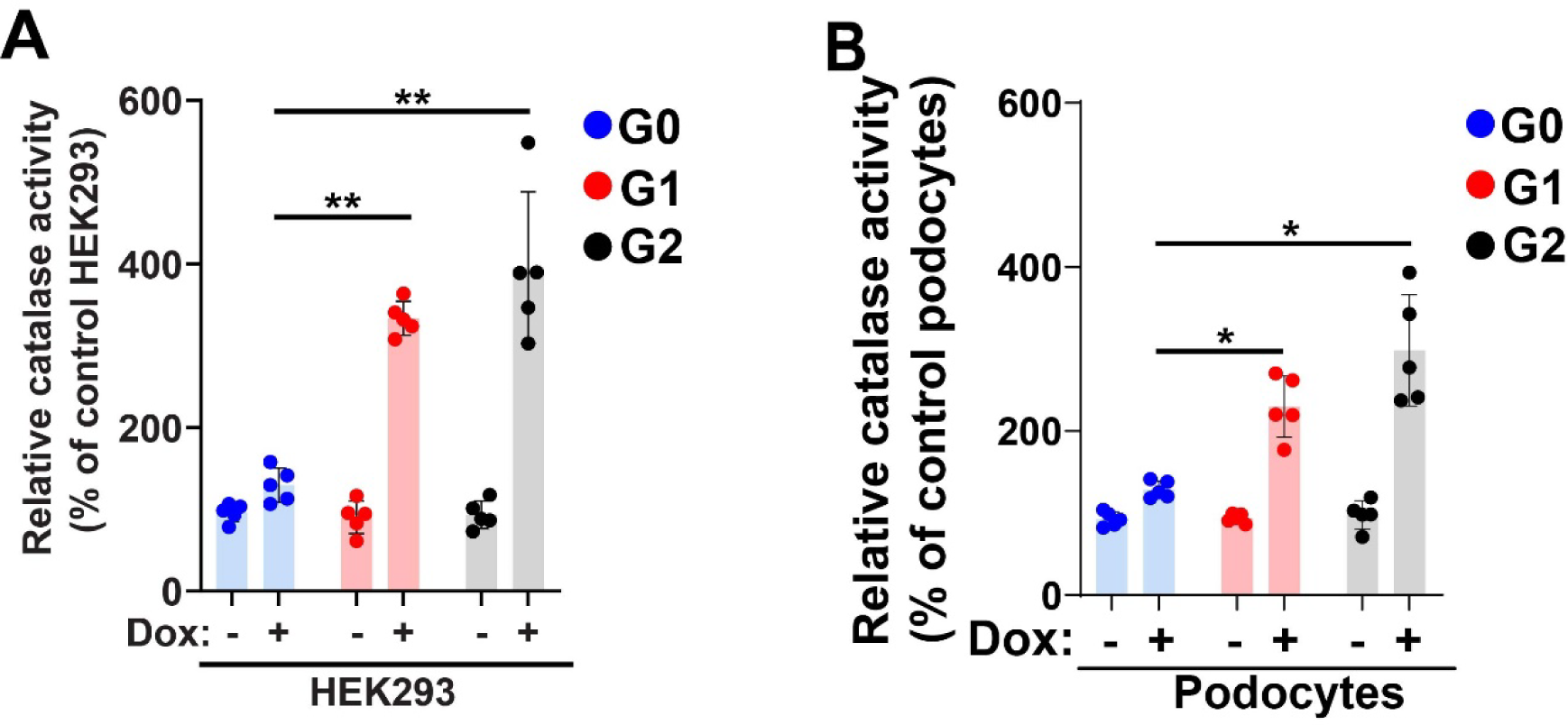
APOL1-G1 or G2 expression results in peroxisomal stress. (**A-B**) Catalase activity is increased in isolated peroxisomes from HEK293 and human podocytes expressing APOL1 G1 and G2 compared to G0 at baseline (in the absence of hypoxia) (n= 5). (**p<0.001, *p<0.05).

**Supplementary Figure 5:**
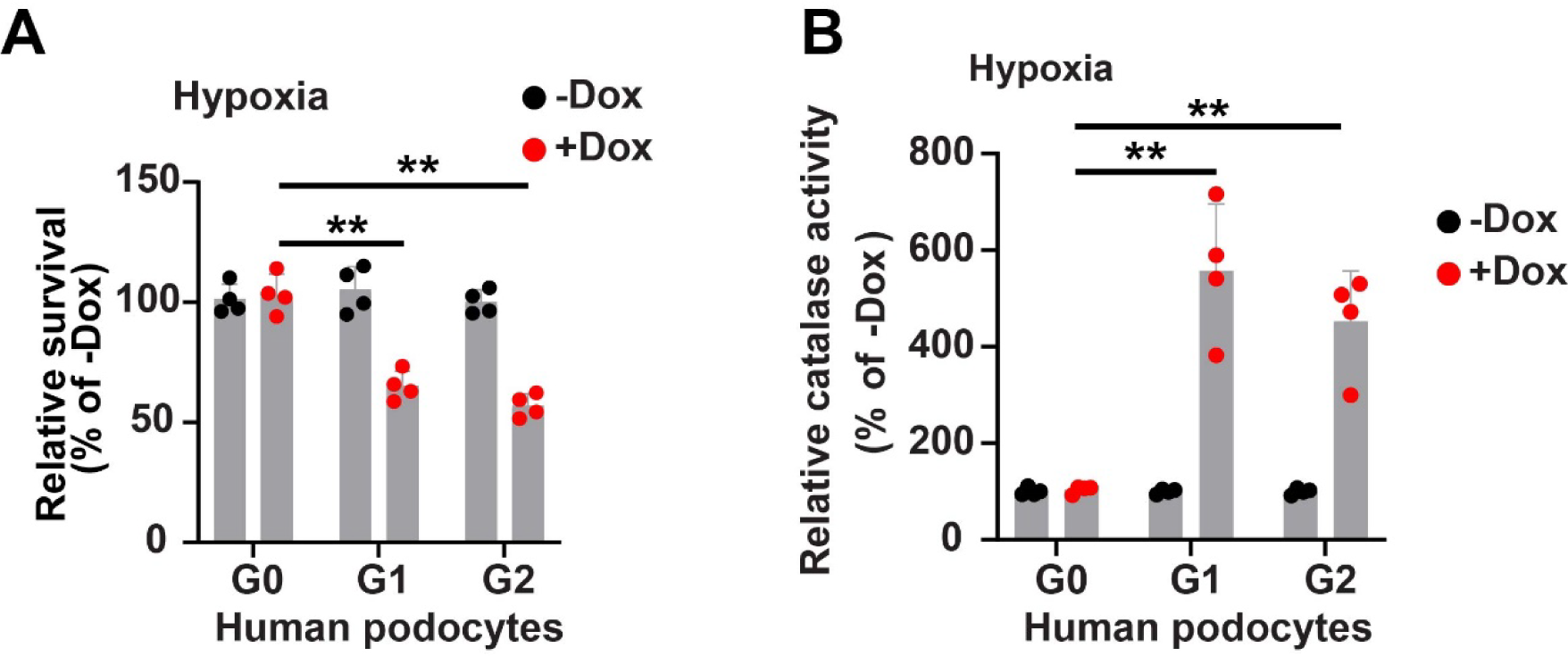
Effect of hypoxia on cytotoxicity and peroxisomal catalase activity in human podocytes expressing APOL1 G0, G1, and G2. (**A**) APOL1-G1 and G2 expression resulted in increased cytotoxicity in human podocytes in hypoxia. (n=4). (**B**) Catalase activity in isolated peroxisomes from human podocytes is higher in G1 and G2 than in G0 in hypoxia. (n=4). (**p<0.001).

**Supplementary Figure 6:**
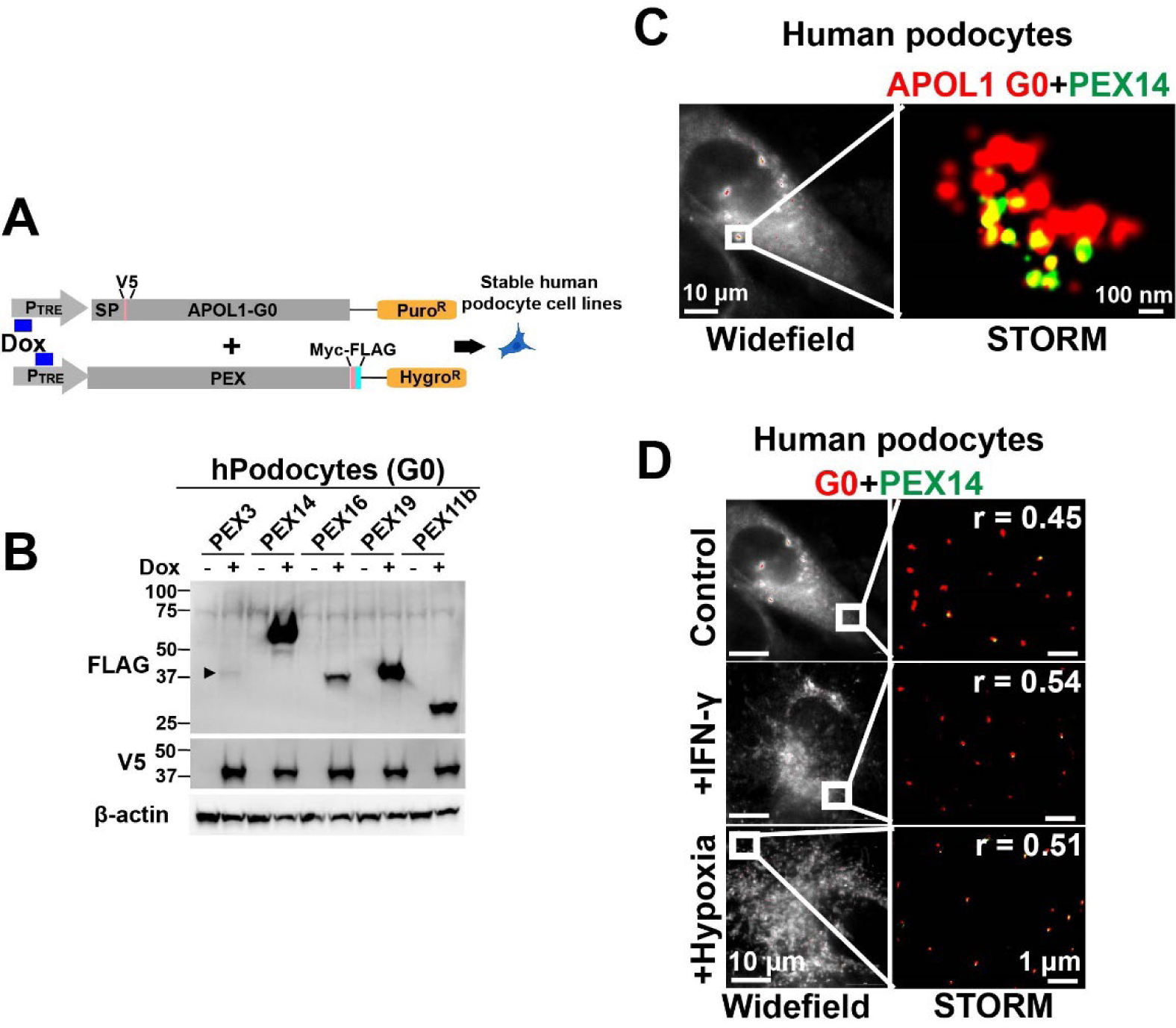
Localization of APOL1 with peroxisomes in human podocytes. (**A**) Schematic of generation of stable doxycycline-inducible human podocytes co-expressing APOL1-G0 with PEX genes. (**B**) Western blot showing cultured human podocytes co-expressing V5 tagged APOL1 G0 with Myc-FLAG tagged PEX proteins under doxycycline control. (**C**) STORM imaging showing colocalization of a fraction of APOL1-G0 (which is predominantly in ER distribution) with peroxisomal membrane protein PEX14 at baseline in human podocytes stably co-expressing doxycycline-inducible APOL1-G0 and PEX14. (**D**) STORM imaging shows colocalization of APOL1-G0 with peroxisomal membrane marker PEX14 in response to IFN-γ, and hypoxia in human podocytes is increased compared to baseline.

**Supplementary Figure 7.**
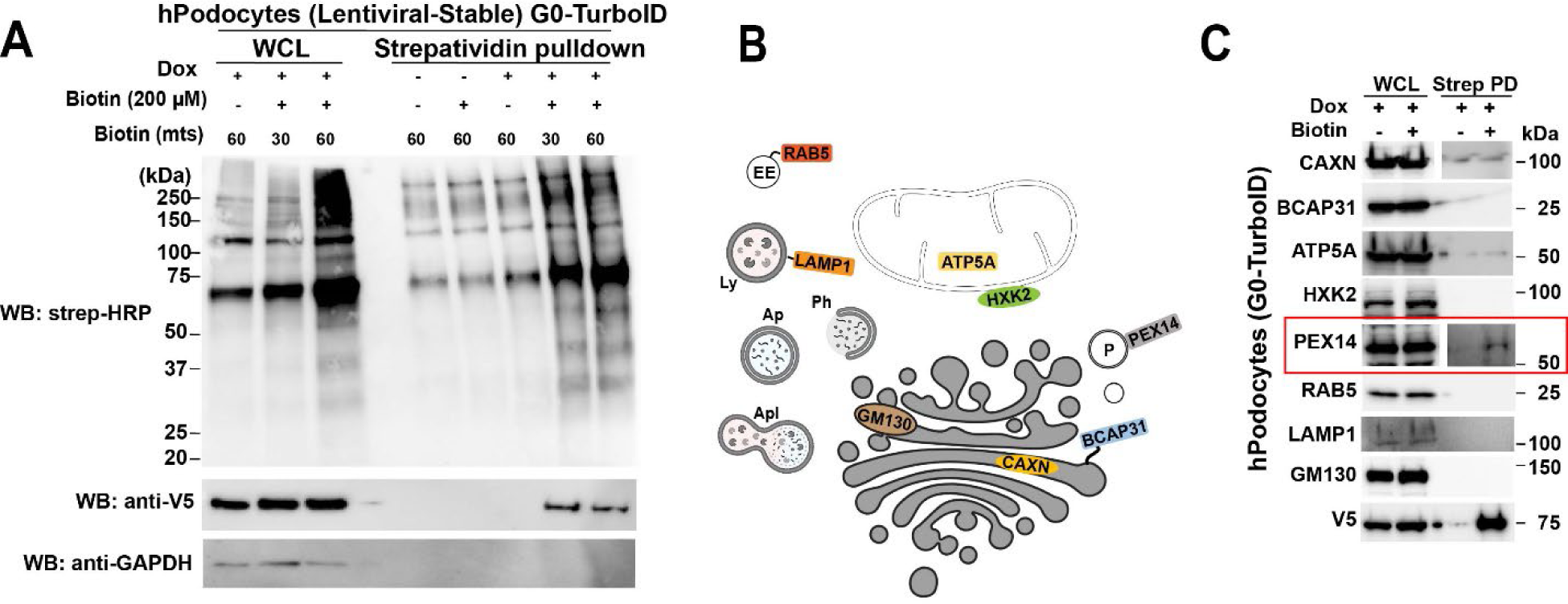
APOL1 is located in proximity to peroxisomes in human podocytes. (**A**) Human podocytes stably expressing doxycycline-inducible G0-TurboID, a biotin ligase, were treated with biotin to activate proximity biotinylation. Western blot shows biotinylated proteins in whole cell lysate and after streptavidin pulldown. (**B-C**) The cellular location and topographical orientation of APOL1 were probed using endogenous organelle markers after proximity biotinylation and pulldown. APOL1-G0-turboID spatially associated with ER matrix (Calnexin), mitochondrial inner membrane/matrix (ATP5A), and peroxisomal membrane (PEX14). No association of APOL1-G0 was observed with ER outer membrane protein (BCAP31), mitochondrial outer membrane (HXK2), Golgi (GM130), early endosome (RAB5), and lysosome (LAMP1).

**Supplementary Figure 8:**
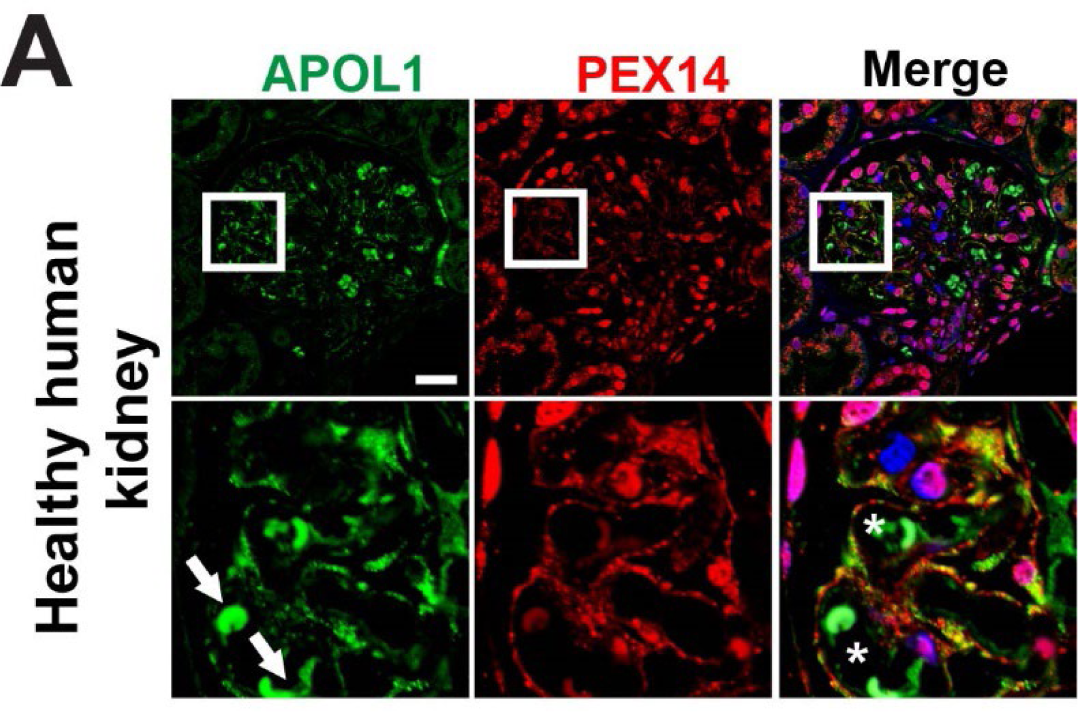
APOL1 localization to peroxisomes in healthy human kidneys. (**A**) Confocal immunofluorescence microscopy showing APOL1 in glomeruli of healthy human kidney colocalizing with peroxisomal membrane protein, PEX14. Arrows denote autofluorescent RBCs and asterisk denote capillary loops. Scale bar: 20 µM.

**Supplementary Figure 9:**
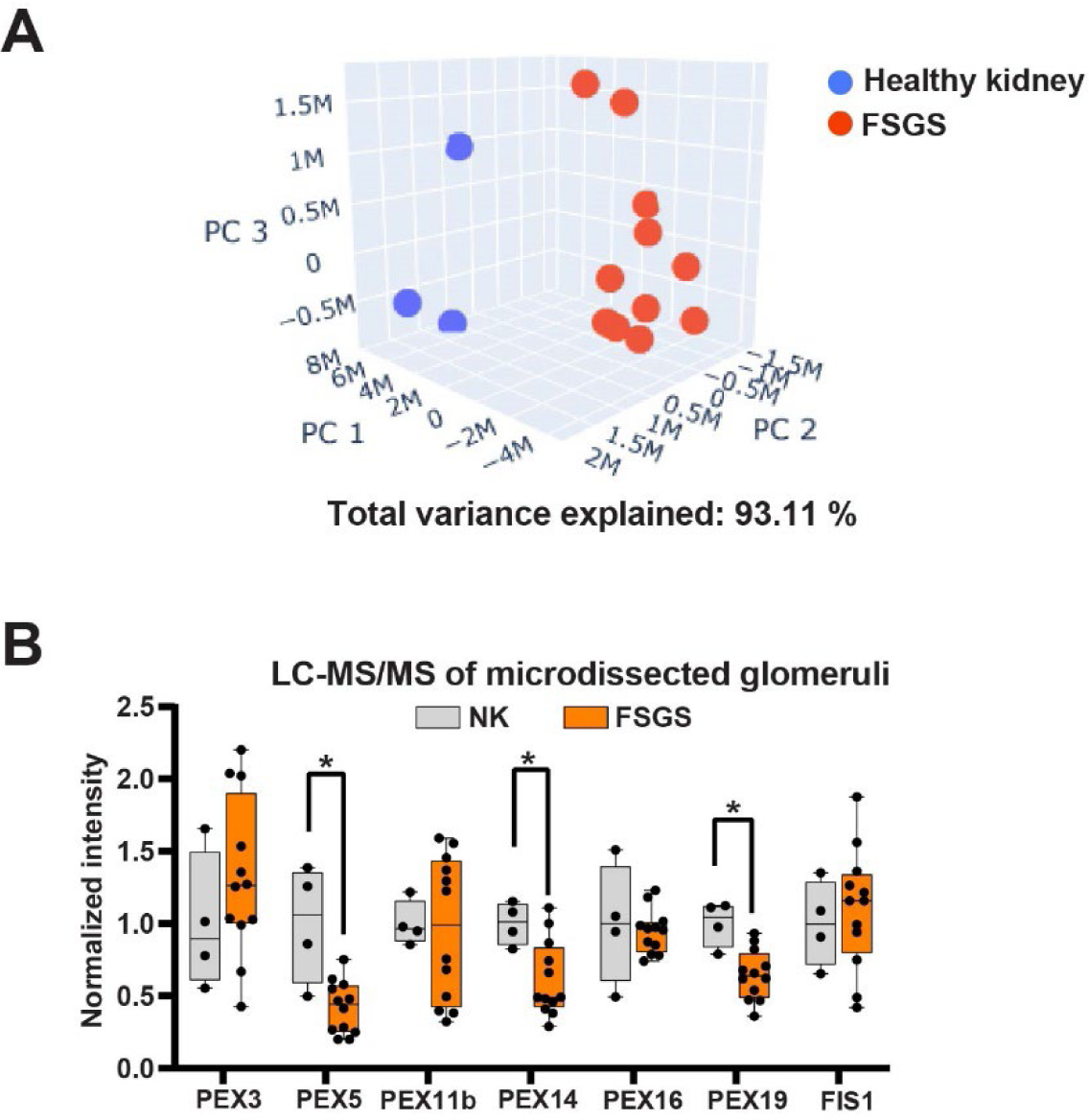
PEX protein expression in FSGS. (**A**) Principal component analysis showing clustering of healthy kidneys and FSGS based on glomerular expression of peroxisomal proteins. Laser capture microdissection and LC-MS of glomeruli were performed in the healthy kidneys (n=3) and FSGS biopsies (n=12). Expression of 231 peroxisomal proteins in glomeruli of healthy kidneys and FSGS were examined from a list curated from previous studies^26,27^ characterizing mammalian peroxisome proteome. (**B**) Laser capture microdissection and LC-MS of glomeruli from healthy dor human kidneys (n=4) and FSGS (n=12) showing expression of PEX proteins. (*p<0.05).

## Supplementary Tables

**Supplementary Table 1:** Plasmids and cell lines used and generated in the study.

| Plasmid | Source | Insert | Inducible (Doxycycline) | Cloning sites | Cell lines |
| --- | --- | --- | --- | --- | --- |
| **pCW57.1** | Addgene (#41393) | *SP (aa 1-30)-V5-APOL1 (aa 31-398) G0 (K150/I228/K255)* | Yes | NheI/Agel | HEK293 and Human podocytes (stable) |
| **pCW57.1** | Addgene (#41393) | *SP (aa 1-30)-V5-APOL1 (aa 31-398) G1 (E150/I228/K255)* | Yes | NheI/Agel | HEK293 and Human podocytes (stable) |
| **pCW57.1** | Addgene (#41393) | *SP (aa 1-30)-V5-APOL1 (aa 31-398) G2 (E150/I228/K255)* | Yes | NheI/Agel | HEK293 and Human podocytes (stable) |
| **pCMCTNT** | Promega | *SP (aa 1-30)-V5-APOL1 (aa 31-398) G0* | No | EcoRI/NotI | HEK293 (transient) |
| **pCMCTNT** | Promega | *SP (aa 1-30)-V5-APOL1 (aa 31-398) G1* | No | EcoRI/NotI | HEK293 (transient) |
| **pCMCTNT** | Promega | *SP (aa 1-30)-V5-APOL1 (aa 31-398) G2* | No | EcoRI/NotI | HEK293 (transient) |
| **pCMCTNT** | Promega | *SP (aa 1-30)-V5-APOL1 (aa 31-398) G0-SVHL* | No | EcoRI/NotI | HEK293 (transient) |
| **pCMCTNT** | Promega | *SP (aa 1-30)-V5-APOL1 (aa 31-398) G1-SVHL* | No | EcoRI/NotI | HEK293 (transient) |
| **pCMCTNT** | Promega | *SP (aa 1-30)-V5-APOL1 (aa 31-398) G2-SVHL* | No | EcoRI/NotI | HEK293 (transient) |
| **pCW57.1** | Addgene (#41393) | *SP (aa 1-30)-V5-APOL1 (aa 31-398)-Linker-turboID* | Yes | NheI/Agel | Human podocytes (stable) |
| **pCW57.1 + pCW57-MCS1-P2A-MCS2** | Addgene | *SP (aa 1-30)-V5-APOL1 (aa 31-398) G0 + PEX3-MYC-FLAG* | Yes | NheI/Agel (for both) | Human podocytes (stable) |
| **pCW57.1 + pCW57-MCS1-P2A-MCS2** | Addgene | *SP (aa 1-30)-V5-APOL1 (aa 31-398) G0 + PEX14-MYC-FLAG* | Yes | NheI/Agel (for both) | Human podocytes (stable) |
| **pCW57.1 + pCW57-MCS1-P2A-MCS2** | Addgene | *SP (aa 1-30)-V5-APOL1 (aa 31-398) G0 + PEX16-MYC-FLAG* | Yes | NheI/Agel (for both) | Human podocytes (stable) |

**Supplementary Table 2:** Details of primary antibodies used in the study.

| **Antibody** | **Vendor** | **Catalog number/Clone ID** | **Source** | **Application** |
| --- | --- | --- | --- | --- |
| **APOL1** | Proteintech | 66124-Ig, clone 1G12D11 | Mouse monoclonal | WB, IF |
| **APOL1** | Genentech | 4.17A5 | Mouse monoclonal | IF |
| **β-tubulin** | Cell signaling | 2128, clone 9F3 | Rabbit monoclonal | WB |
| **GAPDH** | Cell signaling | 2118, clone 14C10 | Rabbit monoclonal | WB |
| **FLAG** | Sigma Aldrich | F1804, clone M2 | Mouse monoclonal | WB |
| **V5** | Cell signaling | 80076, clone E9H8O | Mouse monoclonal | WB, ICC |
| **V5** | Cell signaling | 13202, clone D3H8Q | Rabbit monoclonal | WB |
| **CALNEXIN** | Proteintech | 10427-2-AP | Rabbit polyclonal | WB, ICC |
| **BCAP1** | Proteintech | 11200-1-AP | Rabbit polyclonal | WB |
| **ATP5A** | Abcam | ab14748, clone 15H4C4 | Mouse monoclonal | WB |
| **HXK2** | Abclonal | A0994 | Rabbit polyclonal | WB |
| **RAB5** | Cell Signaling | 3547, clone C8B1 | Rabbit monoclonal | WB |
| **LAMP1** | Abcam | ab25245, clone 1D4B | Rat monoclonal | WB |
| **GM130** | Abcam | ab52649, clone EP892Y | Rabbit monoclonal | WB |
| **PEX14** | Proteintech | 10594-1-AP | Rabbit polyclonal | WB, ICC, IF |
| **PMP70** | Abcam | ab3421 | Rabbit polyclonal | IF |
| **Catalase** | Abcam | ab76110 | Rabbit monoclonal | WB |
| **PDI** | Santa Cruz | sc-74551 | Mouse monoclonal | WB |

Abbreviations- IF: Immunofluorescence (tissue), ICC: Immunocytology, WB: Western blot, IP: Immunoprecipitation

